# A Multi-Criteria Decision Analysis-based Framework for Assessing and Prioritizing Antimicrobial Resistance Risks from Metagenomic Datasets

**DOI:** 10.1101/2025.10.02.25336870

**Authors:** Siddharth Singh Tomar, Krishna Khairnar

## Abstract

Metagenomic sequencing has greatly expanded our ability to detect and characterize resistance genes across diverse environments. However, translating the metagenomic findings into actionable clinical insights remains challenging. We developed a web-based tool that applies a Multi-Criteria Decision Analysis (MCDA) framework to metagenomic AMR datasets. The tool integrates gene abundance with risk attributes derived from the WHO Bacterial Priority Pathogens List, including mortality, transmissibility, and treatability scores. Users can upload AMR detection results alongside customized scoring matrices to generate risk profiles at the samples, species, and drug class levels. Validation was performed using upper respiratory tract samples from SARS-CoV-2 patients and controls in central India, as well as a publicly available dataset from Tanzania. From 48 SARS-CoV-2 samples, 9 produced a total of 10 records involving *Salmonella enterica, Escherichia coli*, and *Streptococcus pneumoniae* across fluoroquinolone, cephalosporin, and macrolide classes (cumulative scores 4.2–146). In contrast, 3 of 48 control samples yielded 3 records, all linked to macrolide resistance in *S. pneumoniae* (scores 5.46–92.43). Analysis of 17 Tanzanian samples identified 2 records, with *Klebsiella pneumoniae* (cephalosporin resistance, score 70.2) and *S. pneumoniae* (macrolide resistance, score 670.02) emerging as priority risks. This framework bridges the gap between raw metagenomic data and clinically relevant risk assessment.

## 1. Introduction

Antimicrobial Resistance (AMR) is a critical public health issue demanding immediate policy and research action. According to the World Health Organisation (WHO), AMR caused 1.27 million global deaths (directly) in 2019 and indirectly contributed to 4.95 million deaths [1]. AMR mitigation requires a multi-pronged approach as resistant pathogens appear across different niches, such as humans, animals, and environmental reservoirs [2]. To maintain the contextual focus on the clinical relevance of AMR, simple, robust and scalable methods or tools are required. Such tools or frameworks are becoming increasingly important for monitoring, evaluating and prioritising the AMR risk across the diverse environments.

Metagenomic sequencing is a technique where the collective genetic material (metagenome) from a sample is sequenced and analysed to characterize the taxonomic composition of organisms present and their functional potential. This technique is particularly useful for detecting and profiling unculturable or novel microbes. With the development and evolution of metagenomics, our ability to detect and characterize AMR genes from diverse niches has increased significantly. While the application of metagenomics in the field of AMR is increasing, the data analysis and interpretation are becoming increasingly complicated [3]. AMR data generated from metagenomic sequencing studies is multi-factorial in nature. Interpretation of Metagenomic AMR data involves a complex interplay of multiple determinants/variables requiring a host-specific and context-aware interpretation framework. Given the multifaceted nature of AMR metagenomics data, it is necessary to investigate the AMR data across multiple criteria. Multi-Criteria Decision Analysis (MCDA) is generally used for dealing with such complex datasets, as it factors in and removes the conflicting and overlapping risk criteria while generating the risk matrix.

Biological and computational researchers are working towards generating robust risk assessment frameworks to better prioritise the AMR risks using the metagenomics data. There are several frameworks that employ MCDA for risk assessment and prioritisation, but these frameworks generally depend on the gene abundance metric and mobility of antibiotic-resistant genes (ARGs) [4,5]. An omics-based approach was used by Zhang et al. (2021), where the researchers used metagenomics data for risk classification (ranking) of already circulating ARGs and predicting the likelihood of currently non-pathogenic ARGs converting into the pathogenic ones through mechanisms like horizontal gene transfer [6]. RESCon is another risk assessment model developed by Martínez et al. (2015). This model ranks ARGs on the basis of their likelihood of circulating among human pathogens. RESCon’s risk attribution algorithm attributes the highest risk scores to ARGs that are more related to the mobile genetic elements. Deep learning algorithms are also being used for developing frameworks for AMR risk assessment. One such tool is DeepARG, developed by Arango-Argoty et al. (2017). DeepARG complements the existing AMR data analysis pipelines for accurate detections of ARGs by reducing false negatives, which is crucial for precise risk assessment [7].

Similarly, PathoFact pipeline concurrently identifies virulence factors, toxins, and AMR genes. It also considers the genomic context, including mobile genetic elements, for more accurate risk assessment [8]. In addition to the pipelines or risk assessment frameworks that come into use post data generation, i.e. during the data analysis step, there are pipelines that could be integrated during the assembly step of the metagenomic data processing. Such assembly-based pipelines can annotate ARGs while identifying their hosts (bacteria); they could also reveal the co-occurrence of ARGs with mobile genetic elements, facilitating the evaluation of environmental risks and the relationship between ARGs, hosts, and environments [9]. These frameworks and tools collectively enhance the accuracy and comprehensiveness of AMR risk assessment, enabling targeted surveillance and intervention strategies to mitigate the spread of high-risk resistance genes.

Despite all the advantages discussed in existing frameworks, these approaches have several limitations. As Many of these tools are context-specific, limiting their generalizability across different ecological or geographical contexts, for example, many existing MCDA tools and risk assessment frameworks are developed and validated within specific environmental or clinical contexts, such as hospital wastewater, urban sewage, or human gut microbiomes. These frameworks may not perform well when applied to other settings like agricultural soils, wildlife reservoirs, or underexplored ecosystems. Another major limitation is the lack of integration with phenotypic resistance data. Additionally, inconsistent annotation of ARGs across different databases leads to redundant and inaccurate results. Despite their influence on ARG spread, current models often overlook environmental and socioeconomic drivers. Many of these existing models are computationally demanding, which limits their utility in resource-poor settings.

To address some of these limitations and provide a user-friendly real-time AMR risk assessment solution, we have developed a tool that implements an MCDA framework on user-provided metagenomic data. This tool allows users to upload two datasets: one with AMR gene detection results (including sample name, drug class, RPM (abundance metric), and detected species/taxa) and another with user-defined MCDA scores across species and drug classes. The MCDA prioritization criteria used for demonstrating this framework were derived from the WHO Bacterial Priority Pathogens List (2024) to make the tool accurate and universally acceptable [10]. Our proposed framework is suitable for being applied to datasets from a clinical background to generate the AMR risk scores in a specific human-health context. Our web-based application implementing this framework will standardise the input datasets into required formats while calculating the species-specific risk scores, considering ARG abundance and MCDA-derived risk factors. This framework takes a “trinity approach” incorporating gene abundance, drug class risk, and species risk to generate cumulative risk scores per sample.

## 2. Materials and Methods

### 2.1. Sample Collection and Metagenomic Analysis

A total of 96 upper respiratory tract samples were analyzed in this study to validate the risk scoring framework. These samples were collected from various districts of central India between March and April 2023 as part of the SARS-CoV-2 genomic surveillance Program. The dataset comprised 48 samples from SARS-CoV-2-positive individuals and 48 samples from a control (SARS-CoV-2-negative) group. All samples underwent Metagenomic next-generation sequencing (mNGS) to characterize the microbiome and resistome composition. The shotgun metagenomic sequencing was done on the NextSeq550 platform.

The raw metagenomic reads were processed using the Chan Zuckerberg ID (CZID) web platform for taxonomic classification and antimicrobial resistance (AMR) gene profiling. CZID’s AMR Pipeline v1.4.2 was used for AMR profiling and annotation [11]. CZID’s AMR Pipeline implements the Resistance Gene Identifier (RGI) tool for aligning the metagenomic reads against the Comprehensive Antibiotic Resistance Database (CARD) [12]. CARD database version 3.2.6 was used in the analysis. The sample-wise output file of combined AMR results contains relevant fields like AMR abundance in reads per million (RPM), read species information, corresponding drug class information, and quality metrics like coverage and depth of the alignment. The results from both the SARS-CoV-2 and Control groups were downloaded as.csv files and then processed separately on the MCDA framework tool to compare the risk profiles of both datasets.

### 2.2. MCDA Risk Matrix Compilation

An MCDA Matrix was designed to evaluate the risk associated with different Bacteria-ARG combinations. The MCDA criteria were selected on the basis of the WHO Bacterial Priority Pathogens List, 2024. Each row in this matrix represented a specific combination of bacterial pathogen and drug class against which the pathogen is resistant, mapped against nine public health-oriented scoring features, the scoring range is from 0.5 (lowest) to 5 (highest). The scoring matrix has the following attributes:

1. **Mortality Score**: Indicates the lethality of infection
2. **Incidence Score:** Prevalence of infection in human populations
3. **Non-fatal Burden Score**: Morbidity and impact on quality of life
4. **Transmissibility Score**: The Rate at which the resistance can transmit
5. **Preventability Score**: Availability of preventive measures
6. **Treatability Score:** Effectiveness of available treatments
7. **Resistance Trend Score**: Indicating whether resistance is increasing over time
8. **Pipeline Score**: Availability of effective drugs in development **(Table 1)**. Additionally, to enhance flexibility and improve the matching of AMR events with scoring data, each row also included alternative names for drug classes commonly used in literature or resistance databases.

**Table 1.** Multi-Criteria Decision Analysis (MCDA) scoring table for computation of the risk scores.

| species | drug_class | mortality_score | incidence_score | non_fatal_burden_score | transmissibility_score | preventability_score | treatability_score | resistance_trend_score | pipeline_score |
| --- | --- | --- | --- | --- | --- | --- | --- | --- | --- |
| *Acinetobacter baumannii* | carbapenem | 3 | 2 | 2 | 1.5 | 2.5 | 3 | 3 | 3 |
| *Pseudomonas aeruginosa* | carbapenem | 3 | 2 | 2 | 2 | 2.5 | 2.5 | 1 | 3 |
| *Klebsiella pneumoniae* | carbapenem | 3 | 2 | 2 | 2.5 | 2.5 | 2.5 | 5 | 3 |
| *Escherichia coli* | carbapenem | 2.5 | 2.5 | 2.5 | 2.5 | 2.5 | 2.5 | 3 | 2 |
| *Enterobacter spp.* | carbapenem | 3 | 1.5 | 1.5 | 1.5 | 2.5 | 2.5 | 4 | 2 |
| *Escherichia coli* | cephalosporin | 2 | 3 | 3 | 3 | 3 | 1.5 | 5 | 2 |
| *Klebsiella pneumoniae* | cephalosporin | 2.5 | 2.5 | 2 | 3 | 3 | 1.5 | 3 | 2 |
| *Proteus spp.* | cephalosporin | 2 | 1.5 | 1.5 | 1.5 | 3 | 1.5 | 2 | 1 |
| *Morganella spp.* | cephalosporin | 1.5 | 0.5 | 0.5 | 1.5 | 3 | 1.5 | 2 | 1 |
| *Citrobacter spp.* | cephalosporin | 2 | 1.5 | 0.5 | 2 | 3 | 1.5 | 2 | 2 |
| *Serratia spp.* | cephalosporin | 2 | 1.5 | 0.5 | 2 | 3 | 1.5 | 2 | 1 |
| *Enterobacter spp.* | cephalosporin | 2 | 2 | 1.5 | 2 | 3 | 1.5 | 2 | 1 |
| *Enterococcus faecium* | vancomycin | 2.5 | 1.5 | 1.5 | 3 | 3 | 1.5 | 3 | 3 |
| *Staphylococcus aureus* | methicillin | 2.5 | 3 | 3 | 3 | 1.5 | 0.5 | 1 | 3 |
| *Salmonella Typhi* | fluoroquinolone | 2 | 1.5 | 2 | 2.5 | 1.5 | 2 | 5 | 3 |
| *Shigella spp.* | fluoroquinolone | 1.5 | 2.5 | 2.5 | 3 | 2 | 1 | 5 | 3 |
| *Salmonella enterica* | fluoroquinolone | 0.5 | 3 | 2.5 | 3 | 3 | 1 | 5 | 2 |
| *Neisseria gonorrhoeae* | cephalosporin | 0.5 | 1.5 | 0.5 | 2 | 2 | 2 | 1 | 3 |
| *Neisseria gonorrhoeae* | fluoroquinolone | 0.5 | 3 | 2 | 2 | 2 | 1.5 | 4 | 3 |
| *Haemophilus influenzae* | ampicillin | 2 | 1.5 | 1.5 | 1.5 | 0.5 | 1 | 2 | 3 |
| *Group A Streptococci* | macrolide | 0.5 | 3 | 2 | 1.5 | 1.5 | 0.5 | 4 | 2 |
| *Group B Streptococci* | penicillin | 1.5 | 0.5 | 0.5 | 0.5 | 1 | 1 | 4 | 2 |
| *Streptococcus pneumoniae* | macrolide | 2 | 2.5 | 3 | 1 | 1 | 0.5 | 2 | 1 |
| *Mycobacterium tuberculosis* | rifampicin | 2.5 | 0.5 | 3 | 2 | 3 | 3 | 3 | 3 |

### 2.3. AMR Risk Scoring and Visualization

Both the AMR Results and MCDA Matrix datasets were uploaded to the tool, and the presence of all essential columns (headers) required for downstream analysis was validated. Species and drug class columns were cleaned and standardized through the removal of trailing descriptors. The case normalization was applied for consistent matching. Instances where multiple microbial species were listed within the `read_species` field were parsed internally and stored into separate columns, thereby enabling species-level granularity and accurate hits in subsequent analyses. To ensure integration between the AMR detection data and the MCDA risk framework, species names were matched to entries in the MCDA matrix via exact string comparison to ensure that the risk scoring only focused on the relevant bacteria-drug combinations. In case no direct match is found in the original drug class column, optional alternative drug class matching was adopted to enhance the sensitivity and accuracy of the matching process. When the relevant bacteria-drug combination from the MCDA matrix finds a direct hit into the AMR dataset or an indirect hit through the alternative drug classes, the tool registers it as a relevant AMR detection event. For each AMR detection event, a quantitative risk score was computed according to the formula:

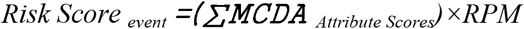

where RPM denotes the normalized abundance of the detected AMR gene in reads per million. AMR detection events for which no valid species–drug class match could be established within the MCDA matrix were excluded from scoring. Risk scores were subsequently aggregated across multiple hierarchical levels, including (i) per sample and drug class, (ii) per sample-species-drug class triplet (Trinity aggregation), and (iii) per species across all samples.

Data visualization was conducted using the `matplotlib` and `seaborn` libraries [13,14], and the complete code was deployed as a dashboard using Streamlit [15]. Visualizations included log-transformed heatmaps depicting AMR risk scores across drug classes and samples, bar plots summarizing total AMR risk per sample, and faceted heatmaps showing both AMR gene abundance (RPM) and cumulative risk across microbial species for each drug class. To enable a clinical context-specific risk profiling, a Trinity-style aggregation approach was employed, where the combination of `sample_name, species`, and `drug_class` served as the analytical unit. For each triplet, the cumulative risk score was calculated as:

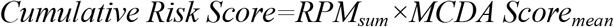

Where *RPM*_*sum*_ represents the total abundance of the relevant AMR gene(s) within the given triplet, and *MCDA Score*_*mean*_ reflects the average of the associated MCDA attribute scores. This generates three outputs in.csv format: (i) a MCDA Data file capturing individual triplet-level metrics, (ii) a Risk Scores Pivot Table summarizing sample-wise risk profiles, and (iii) a Final Aggregated Risk Table providing species- and drug class-level cumulative risk metrics. A detailed workflow of the tool’s execution is shown in **Figure 1**.

**Figure 1.**
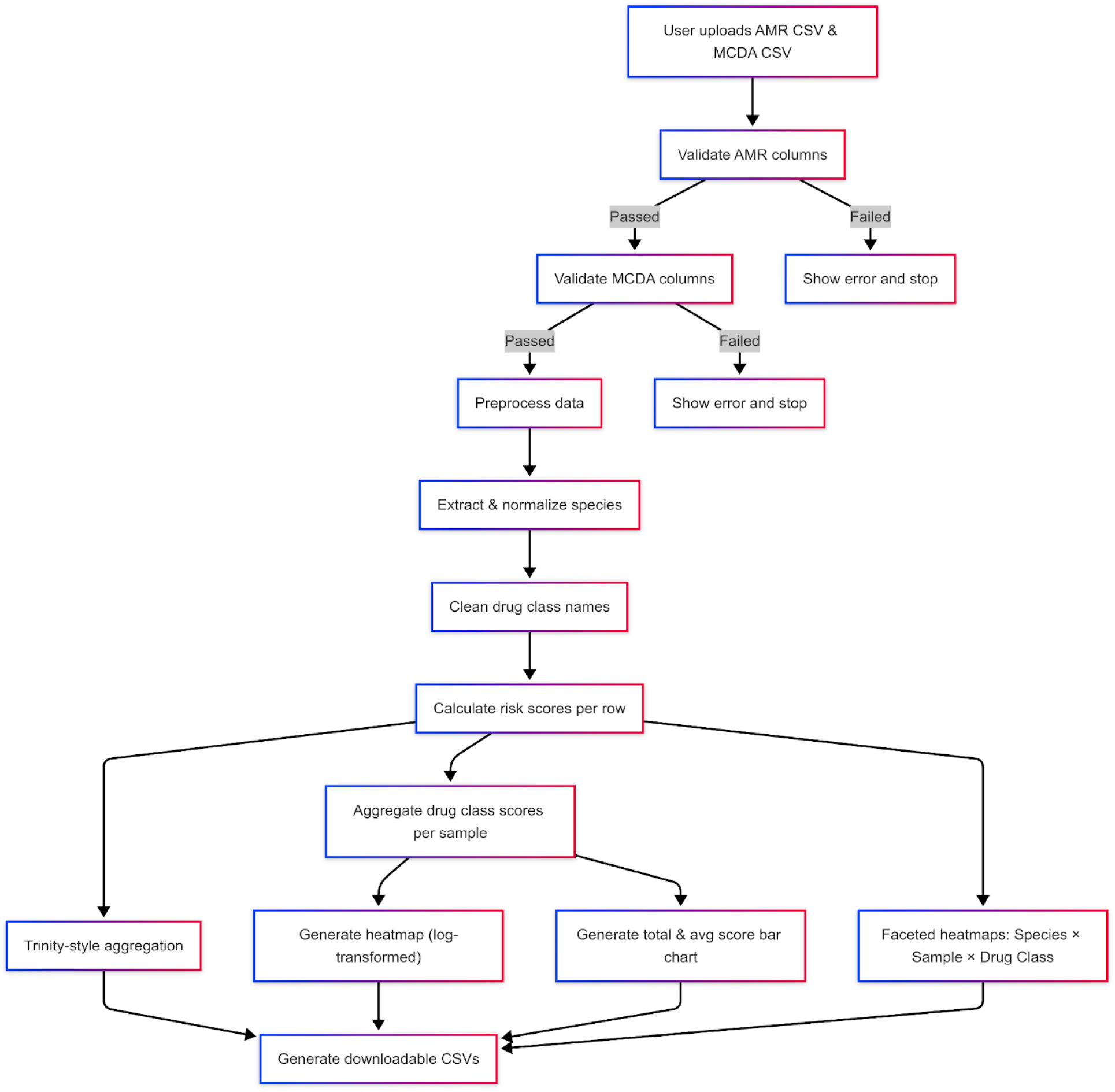
Flowchart showing the execution of the MCDA-AMR tool. The workflow initiates with validation of input datasets, followed by species and drug class normalisation. MCDA is applied to compute record-level and aggregated risk scores. Outputs include log-transformed heatmaps, faceted drug class–species visualizations, bar plots, and CSV files.

## 1. Results

### 3.1. Validation Using In-house Metagenomic Data

The SARS-CoV-2 and control datasets were processed through the MCDA pipeline. Differences were observed in the number of samples yielding AMR risk scores between the two groups. For the SARS-CoV-2 group, out of 48 samples analysed, 9 samples contributed risk score data, yielding a total of 10 records involving three bacterial species: *Salmonella enterica, Escherichia coli*, and *Streptococcus pneumoniae* (**Table 2**). The drug classes represented included fluoroquinolones, cephalosporins, and macrolides. The cumulative risk scores ranged from 4.2 to 146 (**Figure 2)**. For the control group, out of 48 samples analysed, 3 samples contributed risk score data, yielding a total of 3 records, all involving *Streptococcus pneumoniae* and macrolide resistance. The cumulative risk scores ranged from 5.46 to 92.43 **(Table 3)**. The SARS-CoV-2 group exhibited both higher variability and a broader spectrum of AMR risks across antimicrobial classes compared to the control group, which showed a more restricted AMR risk profile **(Figure 3)**.

**Table 2.** Detailed MCDA table for SARS-CoV-2 samples data.

| sample_name | species | drug_class | mortality_score | incidence_score | non_fatal_burden_score | transmissibility_score | preventability_score | treatability_score | resistance_trend_score | pipeline_score | rpm | mcda_score | cumulative_risk_score |
| --- | --- | --- | --- | --- | --- | --- | --- | --- | --- | --- | --- | --- | --- |
| 23G214-5G-270_S7 | *salmonella enterica* | fluoroquinolone | 0.5 | 3 | 2.5 | 3 | 3 | 1 | 5 | 2 | 7.3 | 20 | 14 |
| 23G214-5G-271_S8 | *salmonella enterica* | fluoroquinolone | 0.5 | 3 | 2.5 | 3 | 3 | 1 | 5 | 2 | 1.59 | 20 | 31. |
| 23G214-5G-277_S14 | *salmonella enterica* | fluoroquinolone | 0.5 | 3 | 2.5 | 3 | 3 | 1 | 5 | 2 | 5.96 | 20 | 119. |
| 23G214-5G-279_S16 | *salmonella enterica* | fluoroquinolone | 0.5 | 3 | 2.5 | 3 | 3 | 1 | 5 | 2 | 1.12 | 20 | 22. |
| 23G214-5G-281_S18 | *salmonella enterica* | fluoroquinolone | 0.5 | 3 | 2.5 | 3 | 3 | 1 | 5 | 2 | 0.21 | 20 | 4. |
| 23G214-5G-294_S31 | *salmonella enterica* | fluoroquinolone | 0.5 | 3 | 2.5 | 3 | 3 | 1 | 5 | 2 | 3.15 | 20 | 6 |
| 23G214-5G-298_S35 | *escherichia coli* | cephalosporin | 2 | 3 | 3 | 3 | 3 | 1.5 | 5 | 2 | 0.77 | 22.5 | 17.32 |
| 23G214-5G-298_S35 | *streptococcus pneumoniae* | macrolide | 2 | 2.5 | 3 | 1 | 1 | 0.5 | 2 | 1 | 0.77 | 13 | 10.0 |
| 23G214-5G-303_S40 | *streptococcus pneumoniae* | macrolide | 2 | 2.5 | 3 | 1 | 1 | 0.5 | 2 | 1 | 8.98 | 13 | 116.7 |
| 23G214-5G-305_S42 | *escherichia coli* | cephalosporin | 2 | 3 | 3 | 3 | 3 | 1.5 | 5 | 2 | 3.16 | 22.5 | 71. |

**Table 3.**
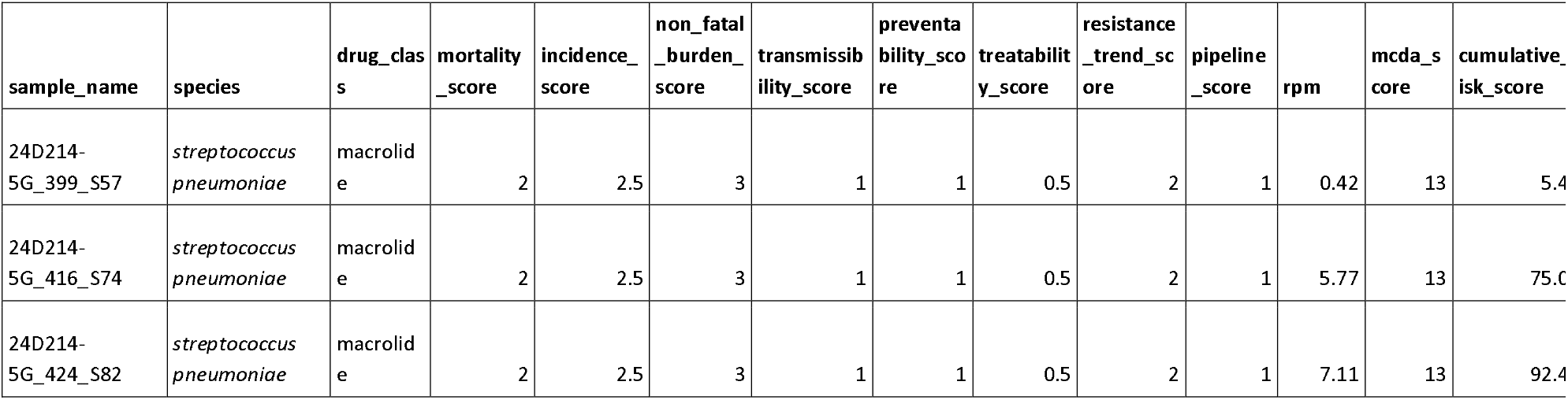
Detailed MCDA table for Control samples data.

| sample_name | species | drug_classes | mortality_score | incidence_score | non_fatal_burden_score | transmissibility_score | preventability_score | treatability_score | resistance_trend_score | pipeline_score | rpm | mcda_score | cumulative_risk_score |
| --- | --- | --- | --- | --- | --- | --- | --- | --- | --- | --- | --- | --- | --- |
| 24D214-5G_399_S57 | *streptococcus pneumoniae* | macrolide | 2 | 2.5 | 3 | 1 | 1 | 0.5 | 2 | 1 | 0.42 | 13 | 5.4 |
| 24D214-5G_416_S74 | *streptococcus pneumoniae* | macrolide | 2 | 2.5 | 3 | 1 | 1 | 0.5 | 2 | 1 | 5.77 | 13 | 75.0 |
| 24D214-5G_424_S82 | *streptococcus pneumoniae* | macrolide | 2 | 2.5 | 3 | 1 | 1 | 0.5 | 2 | 1 | 7.11 | 13 | 92.4 |

**Figure 2:**
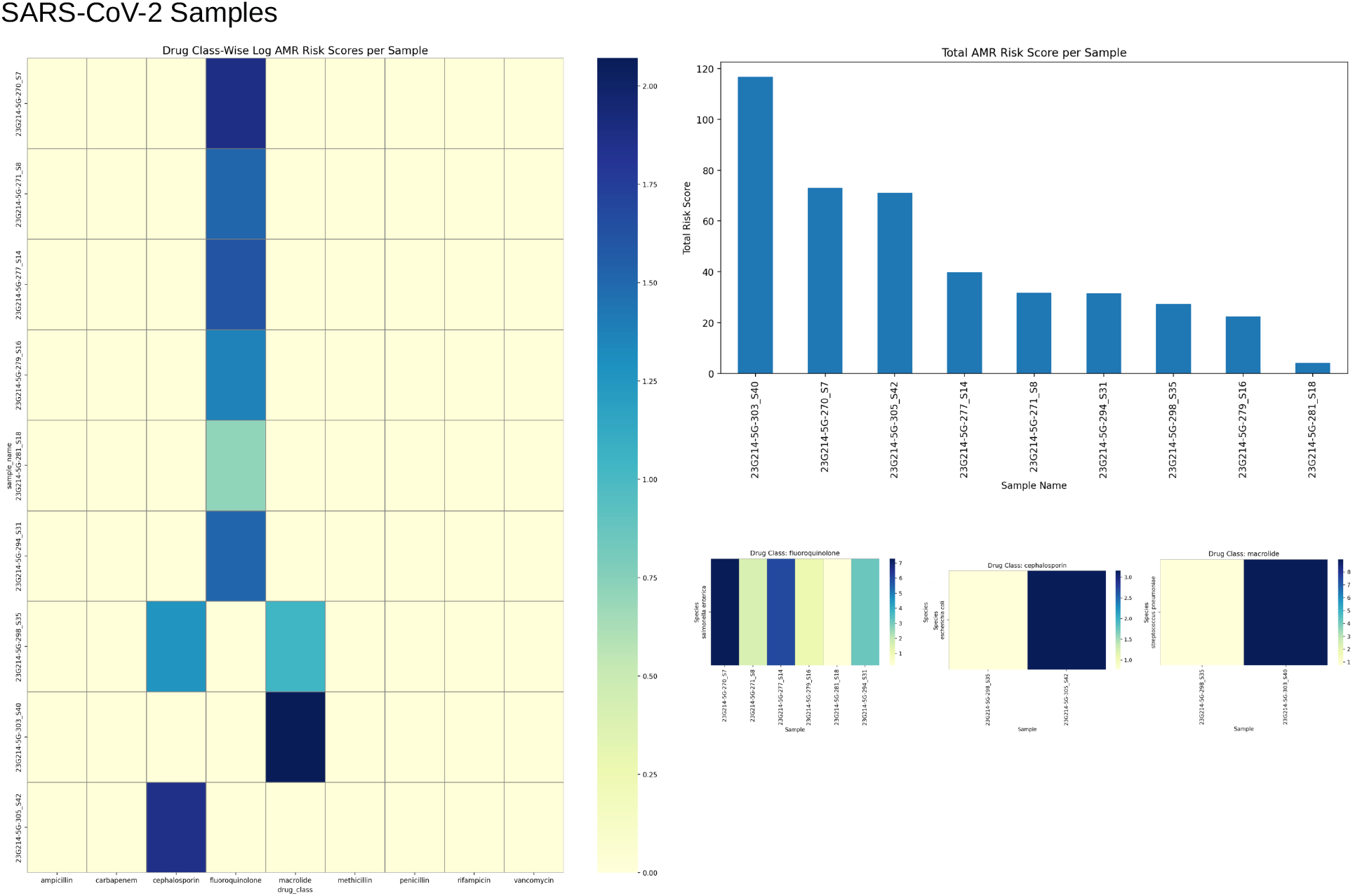
Antimicrobial resistance (AMR) risk scores in SARS-CoV-2 samples. The heatmap (left) depicts log-transformed AMR risk scores across different antibiotic drug classes for each SARS-CoV-2 sample. The bar chart (top right) shows the total AMR risk score aggregated per sample, highlighting inter-sample variability.

**Figure 3:**
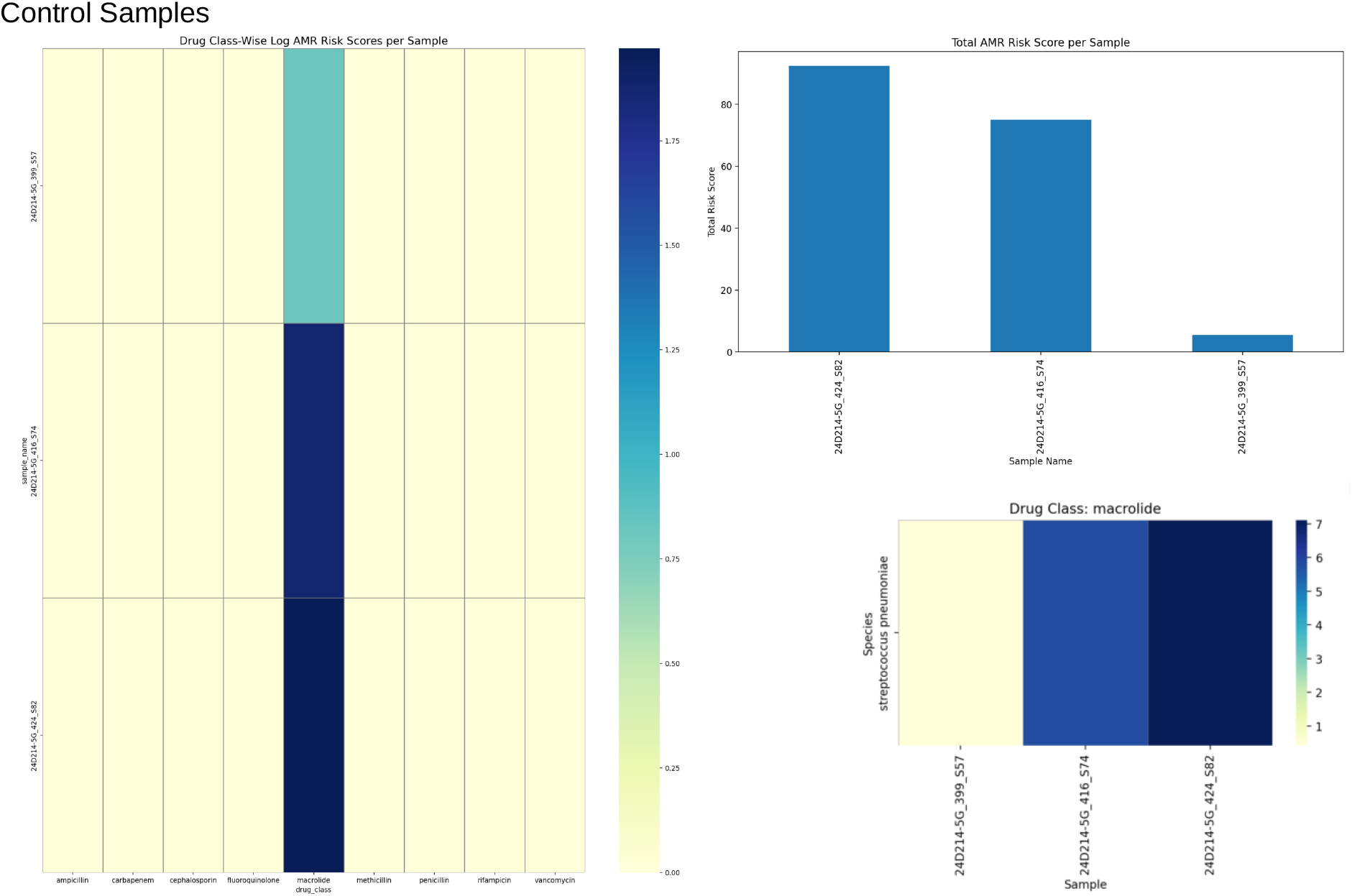
Antimicrobial resistance (AMR) risk scores in Control samples. The heatmap (left) depicts log-transformed AMR risk scores across different antibiotic drug classes for each control sample. The bar chart (top right) shows the total AMR risk score aggregated per sample, highlighting inter-sample variability.

### 3.2. Validation Using Open-source Metagenomic Data

A publicly available dataset from Tanzania, ***“ARI-Mazoezi”***, published on CZID by Emelesiana Magulu (2025), was used to validate the MCDA pipeline. The data for 17 samples were analysed through the pipeline, out of which two samples contributed risk score data, yielding a total of two records involving *Klebsiella pneumoniae* and *Streptococcus pneumoniae* **(Table 4)**. The drug classes represented included cephalosporins and macrolides. The cumulative risk scores were 70.2 and 670.02, respectively, while the corresponding average risk scores were 7.8 and 74.45. The visualisation of these results is shown in **Figure 4**. These findings highlight species-specific differences in antimicrobial resistance risk profiles, with macrolide resistance in *Streptococcus pneumoniae* contributing the highest cumulative burden.

**Table 4.** Detailed MCDA table for open source samples data.

| sample_name | species | drug_class | mortality_score | incidence_score | non_fatal_burden_score | transmissibility_score | preventability_score | treatability_score | resistance_trend_score | pipeline_score | rpm | mcda_score | cumulative_risk_score |
| --- | --- | --- | --- | --- | --- | --- | --- | --- | --- | --- | --- | --- | --- |
| ARH-4077_S17 | *klebsiella pneumoniae* | cephalosporin | 2.5 | 2.5 | 2 | 3 | 3 | 1.5 | 3 | 2 | 3.6 | 19.5 | 70.2 |
| KDH-7035_S5 | *streptococcus pneumoniae* | macrolide | 2 | 2.5 | 3 | 1 | 1 | 0.5 | 2 | 1 | 51.54 | 13 | 670.02 |

**Figure 4:**
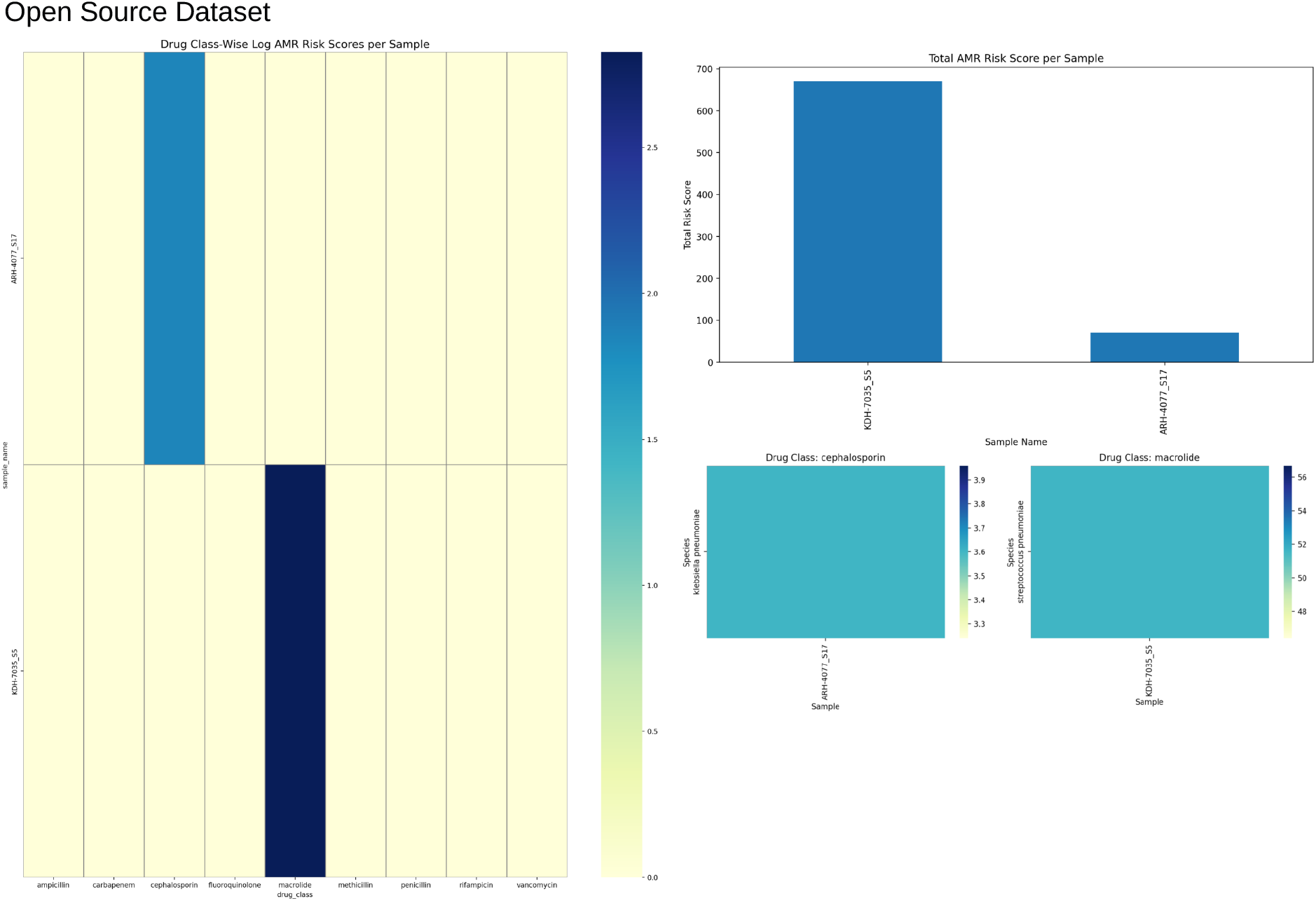
Antimicrobial resistance (AMR) risk scores in the open source data samples. The heatmap (left) depicts log-transformed AMR risk scores across different antibiotic drug classes for each sample. The bar chart (top right) shows the total AMR risk score aggregated per sample, highlighting inter-sample variability.

## 2. Discussion

The results demonstrate the utility of the proposed MCDA-based framework in translating metagenomic data into actionable AMR risk profiles. The results from the in-house dataset revealed differences in AMR risk distribution between SARS-CoV-2 and control samples. The SARS-CoV-2 group yielded a larger number of records. This finding aligns with the recent reports indicating that the SARS-CoV-2 infection leads to significant disruption of the host microbiome and increases the abundance and diversity of antimicrobial resistance genes, especially in the respiratory tract [16,17,18]. These changes are associated with disease severity and may be exacerbated by antibiotic use during COVID-19 management. While the control samples generated fewer risk scores, limited to *Streptococcus pneumoniae* and macrolide resistance, this is indicative of a limited resistance landscape. These findings illustrate the framework’s sensitivity to both pathogen diversity and drug-class specificity; this feature is not commonly captured by traditional abundance-only approaches.

We have applied the framework to an external metagenomic dataset from Tanzania. Among 17 samples, two yielded AMR risk scores, identifying *Klebsiella pneumoniae* and *Streptococcus pneumoniae* as priority species. Notably, macrolide resistance in *S. pneumoniae* accounted for the highest cumulative burden, consistent with global trends recognizing macrolide resistance in *S. pneumoniae* as a clinically important and treatment-limiting factor. Resistance rates range from <10% to >90% depending on geography, with particularly high resistance rates in parts of Asia, Africa and Latin America [19,20,21].

The results demonstrate that integrating multiple decision criteria into risk assessment beyond the traditional abundance-based ranking helps in better contextualisation of metagenomic results for making an informed clinical decision. For example, in the open-source dataset, *S. pneumoniae* macrolide resistance scored disproportionately high in cumulative burden due to its clinical importance, not just its relative abundance. This highlights the added interpretability and clinical relevance the MCDA framework introduces by bridging the gap between raw metagenomic outputs and public health priorities.

The framework’s ability to differentiate among the datasets on the basis of their clinical origin and to identify species and drug-class-specific risks demonstrates its potential use case in AMR surveillance programs, particularly in clinical settings. While the validation on limited datasets underscores proof-of-concept, larger-scale applications and validation across diverse datasets, such as data originating from Environmental and community-level surveillance, are required to fully test scalability and generalizability. Integration of these results with phenotypic resistance data and correlation with clinical outcomes remains an important step to further strengthen accuracy and reduce redundancy.

## Conclusion

The results indicate that the MCDA framework offers a scalable, context-aware, and clinically relevant solution for AMR risk assessment from metagenomic data. This framework aligns the metagenomics outputs with WHO prioritization criteria to generate interpretable visualizations. This tool could serve as a bridge between complex metagenomics datasets and actionable insights for policy, surveillance, and clinical decision-making.

## Supporting information

The data is available as supplementary data with the file name combined supplementary data

## Declaration of Copyright

The tool has been granted copyright registration by the Registrar of Copyrights, India. **(Diary No: SW-31224/2025-CO and ROC number SW-2025021576)**.

## Data availability statement

The data is available as supplementary data with the file name **combined supplementary data** Any additional data, if required, will be made available on request by the authors. The GUI tool implementing the framework is published as an open source app on Streamlit and could be accessed through the following URL: https://mcda-amr-risk-assessment-pipeline-eepm-neeri.streamlit.app/

## Author declaration

The authors assure that the research has followed all ethical guidelines and received approvals from the Institutional Ethics Committee for Research on Human Subjects (IEC) of CSIR-NEERI, Nagpur-20, India. **(<u>Reference No.: Eth.Com./001/IEC/02/2023</u>**) Necessary consent from patients/participants has been obtained, and relevant institutional documentation has been archived. This manuscript is approved by the institutional Knowledge Resource Center (KRC) of CSIR-NEERI, bearing KRC No. CSIR-NEERI/KRC/2025/SEP/EEPM/1

## Confidentiality declaration

Sample IDs are masked IDs and cannot be traced to participant details.

## Author contribution statement

SST and KK have contributed equally to the conceptualization, experimentation, and data analysis of this study.

## Conflict of interest statement

The authors declare no conflict of interest

## Acknowledgement

The authors are thankful to CSIR-NEERI for providing funds under project OLP-57 (March 2023 -April 2024) for conducting this study

## Notes

### Competing Interest Statement

The authors have declared no competing interest.

### Author Declarations

The authors assure that the research has followed all ethical guidelines and received approvals from the Institutional Ethics Committee for Research on Human Subjects (IEC) of CSIR-NEERI, Nagpur-20, India. (Reference No.: Eth.Com./001/IEC/02/2023) Necessary consent from patients/participants has been obtained, and relevant institutional documentation has been archived.

